# Modelling Capacity on London Underground Carriages in Light of UK Government Mandated Social Distancing Rules in Response to the Covid-19 Pandemic

**DOI:** 10.1101/2020.05.26.20113019

**Authors:** Justin Marley

**Affiliations:** Kingswood Centre, Essex Partnership University NHS Foundation Trust, CO4 5JY

**Keywords:** COVID19, SARS-CoV-2, social distancing, London Underground, London transport

## Abstract

The UK has been affected by the COVID-19 pandemic and London in particular has experienced a large number of cases. The London Underground is a key part of the transport for London. The UK Government has implemented social distancing rules meaning that people should be 2 metres from each other. The current paper models the impact of the social distancing on the carrying capacity of 10 different underground and overground carriages. The model determines the optimal standing and seating capacity for the different carriages and identifies logistical approaches to the seating and standing arrangements.

## Introduction

A novel coronavirus SARS-COV2 causing Pneumonia was identified in Wuhan, China in 2019 (Zhou et al, 2020). The illness resulting from the virus was named COVID-19. The COVID-19 outbreak was reported to the World Health Organisation on December 31^st^ 2019 (WHO, 2020). The outbreak later developed into a pandemic. The United Kingdom was affected and London in particular has experienced a significant proportion of cases of COVID-19 and deaths (The London Intelligence, 2020). The UK Government has implemented a phased approach to managing the pandemic with the initiation of a lockdown.

Essential workers in London have been travelling to work. An important aspect of this in London has been the use of the London underground when alternative modes of transport are not possible. There is evidence that the London Underground can be a source of transmission of SARS-COV2. In one study, the researchers found an increased rate of infection in London boroughs with access to an underground interchange (Sasidharan et al, 2020). At one point, it was reported that 30% of selected staff groups at Transport for London were unable to come to work as they were either self-isolating or ill with COVID-19 (Transport for London, 2020).

The current paper aims to examine what impact the current UK Government mandated social distancing policy has on the capacity of London Underground cars (including overground cars).

## Methodology

The modelling was done in two parts. Firstly a literature search identified relevant data. Secondly the data was used to inform the calculations to derive the carrying capacity and these calculations are further outlined below.

## Literature Search

A search of the scientific literature was undertaken using Pubmed (PubMed, 2020), MedRxiv (MedRxiv, 2020) and BioRxiv (BioRxiv, 2020) using the following search terms: ‘London underground and Covid-19’, ‘underground capacity and Covid-19’, ‘underground and social distancing’. A supplementary search was undertaken using Google using the search term ‘London underground capacity’ to obtain technical information about the underground transport.

## Calculating Generic Standing Capacity

Standing capacity was modelled using social distancing rules. A person must stand 2 metres from other people. This is illustrated in figure 1.

**Figure 1:**
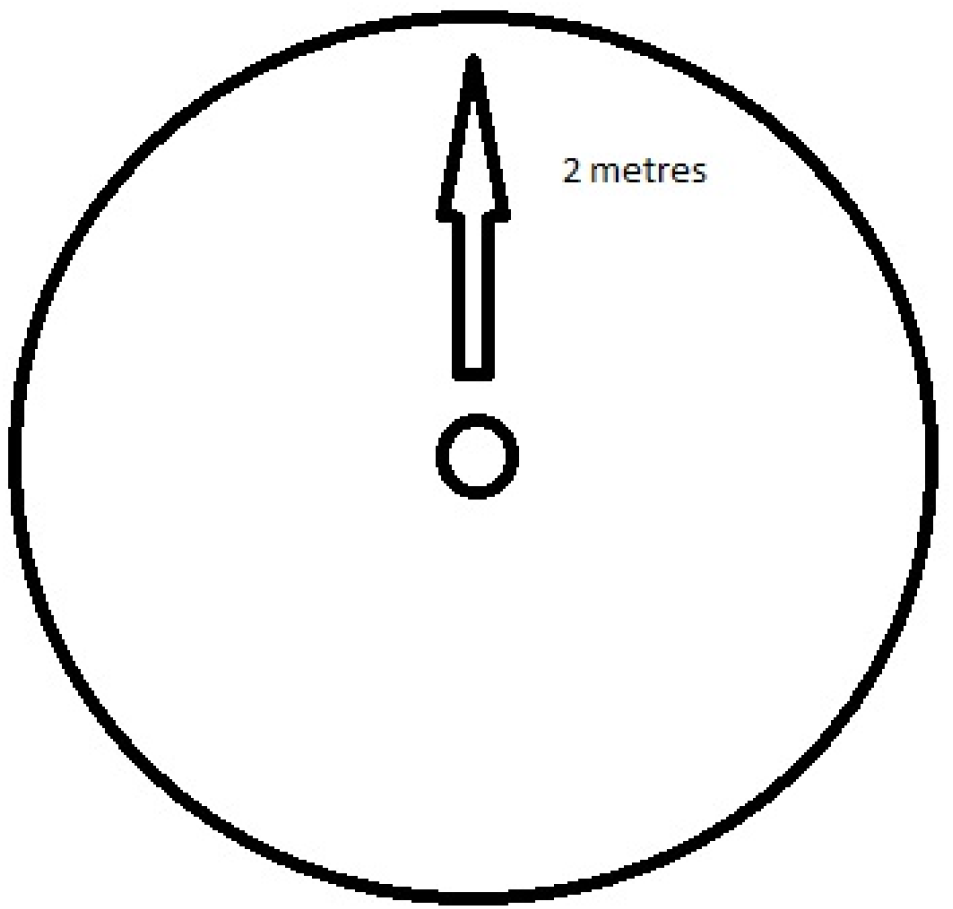
Illustrating a person standing 2 metres from others.

The distance of 2 metres thus forms the radius of a circle. The area of the circle can be calculated from

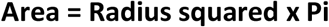

Therefore the area that one person would occupy whilst adhering to UK social distance policy is

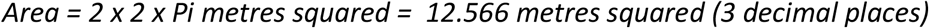

Translating this into density, the current UK social distancing policy requires a person density of

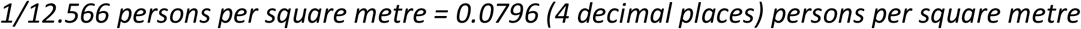

Data was utilised from a website (Ian Visits, 2020) in which underground train capacity was derived from other data sources (What do they know, 2020(a))(What do they know, 2020(b)). A figure of 5 persons-per-metre squared which can be seen in the included Rolling Stock Data Sheet (What do they know, 2020 (a)) was used to populate a table on the site (Ian Visits, 2020) which included standing and sitting capacities. Therefore the initial transformation of standing capacity when applied to this data would result in a 62.8-fold reduction in standing capacity before other factors are applied.

## Estimating Seating Capacity

Retrieved data was used to estimate seating capacity. Microsoft Paint was used to overlay a grid on the schematics and enable measurements. The resulting measurements were used in the analysis of the impact of social distancing on seating and standing capacity.

## Modifying Standard Capacity and Estimating Final Capacity

Retrieved data was used to estimate the impact of social distancing on the interaction between sitting and standing capacities and used to derive a final capacity. This was done using the individual schematics for each subtype of London underground cars.

## Results

### Literature Search

The literature search was completed on 25.5.20 across 3 databases and 1390 papers were identified and are further outlined in Table 1. Of these, only 2 were relevant to the present paper but did not provide any data for the purposes of modelling. A Google search identified a website citing data for carriage capacity based on a previously retrieved Freedom of Information request.

**Table 1.** Number of articles identified in literature search for each database and search term

| Search Terms | Pub Med | MedRxiv | BioRxiv |
| --- | --- | --- | --- |
| 'London underground and Covid-19' | 2 | 7 | 1 |
| 'underground capacity and Covid-19' | 0 | 1052 | 229 |
| 'underground and social distancing' | 0 | 10 | 89 |

### Applying the Reduced Standing Density Secondary to Social Distancing Policy

The results of the first transformation are shown in Table 2

**Table 2:**
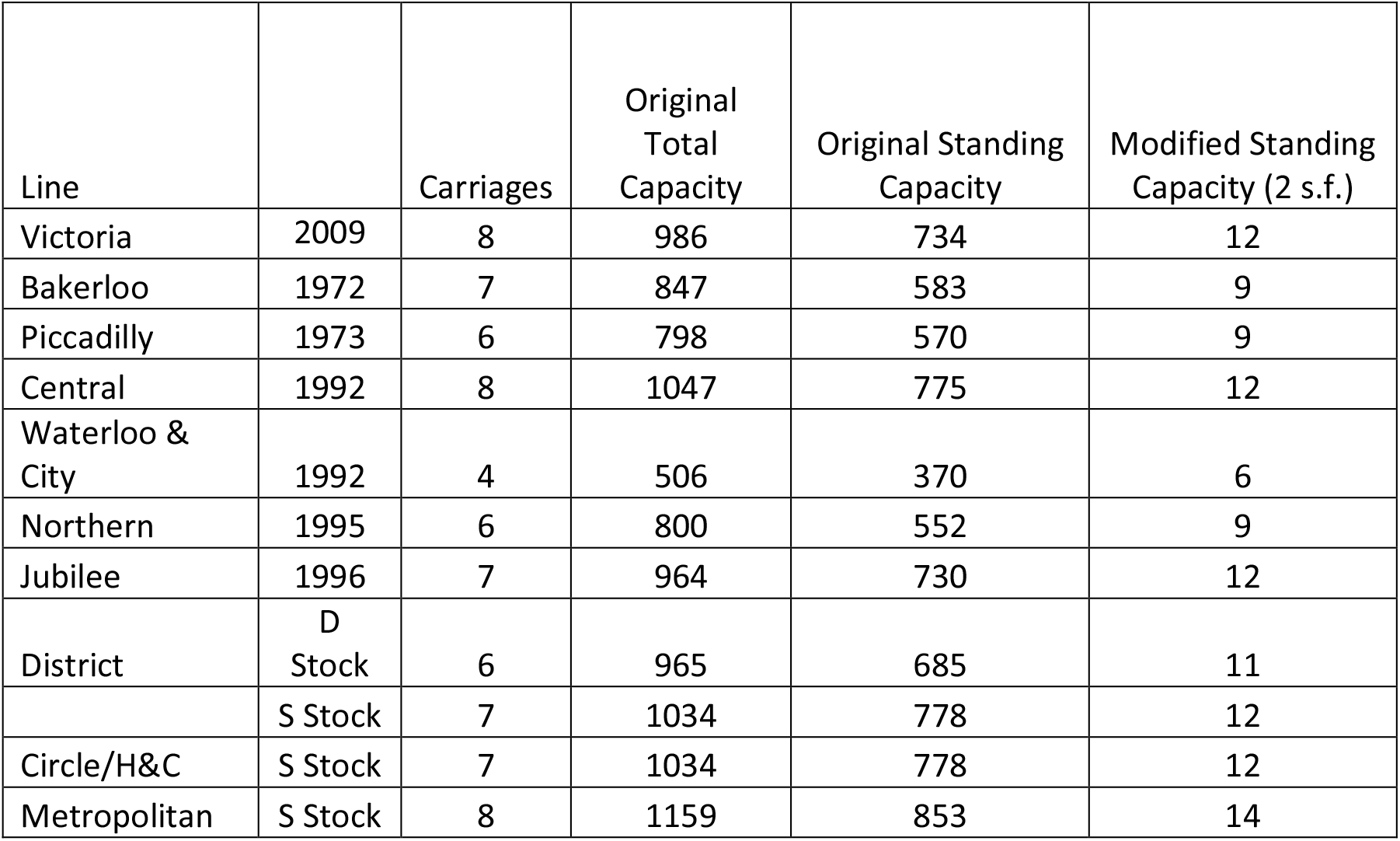
Effect of Social Distancing on Carriage Standing Capacity Without Reference to Sitting Capacity

### Estimating Seating Capacity From a Worked Example

The 1967 Victoria Line tube stock schematics for the trailer car were used to derive seating capacity (WDTK, 2020). The grid overlay is shown in figure 2.

**Figure 2.**
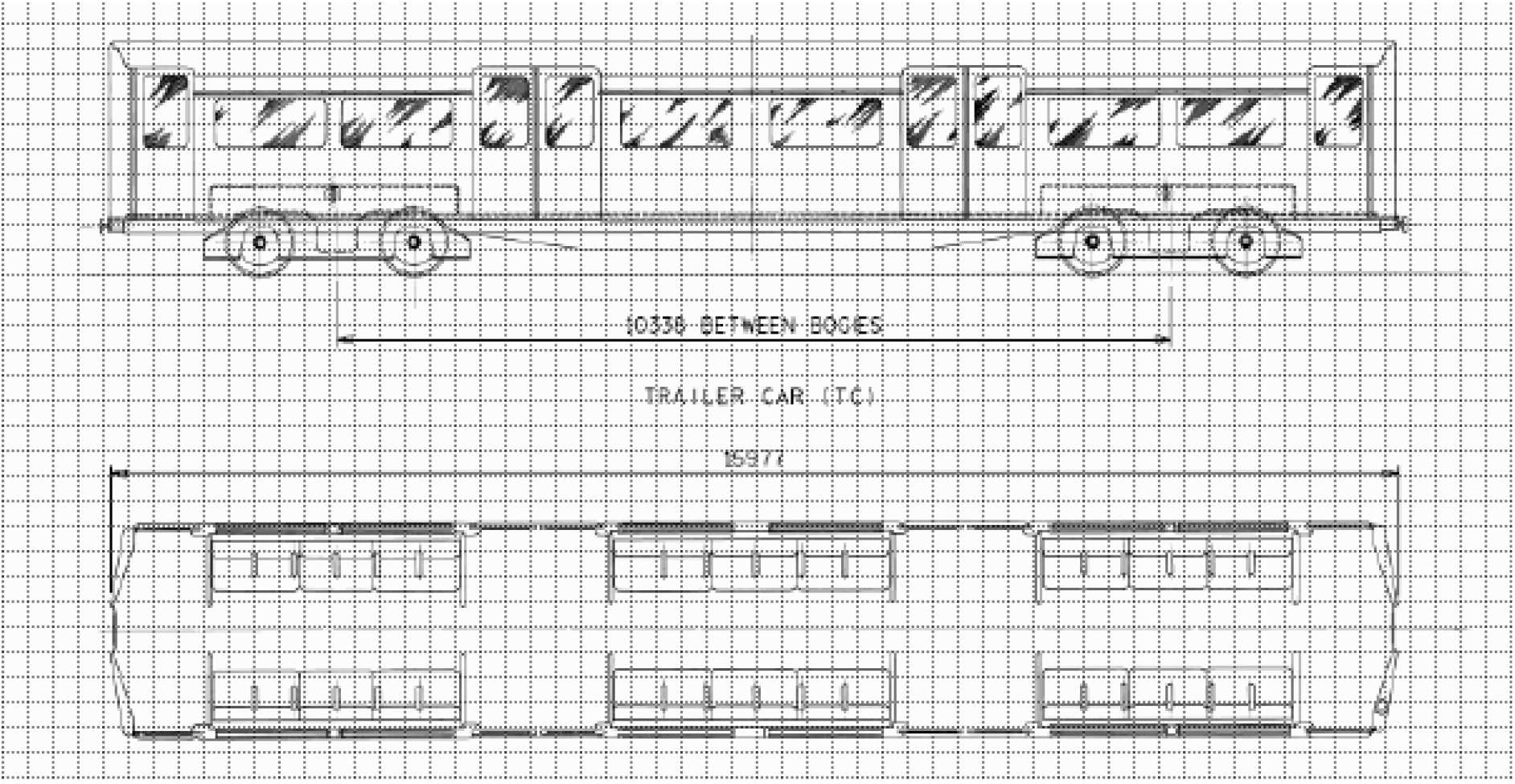
Grid Overlay on 1967 Victoria Line Schematics for Trailer Car

### Worked Example

The measurements on the schematics indicate that the trailer car length is 54 ft 5 inches (16.58 metres (2DP)). The length of the carriage is 50 squares on the superimposed grid and therefore each square is approximately 33cm. The carriage contains 3 groups of 12 seats. However 2 metres is equivalent to the width of 6 squares. Therefore any seating in the same row must accommodate a separation of 6 squares and therefore a maximum of 2 people per row. The distance from the back of one seat to the back of the seat on the opposing row is 7 squares and therefore the seating arrangements on opposing rows can be independent of each other. The total seating capacity is reduced from 36 per carriage in this case, to 12 representing 33% of total seating capacity. These findings are generalised to the other types of carriage to derive a total seating capacity as shown in Table 3.

**Table 3:**
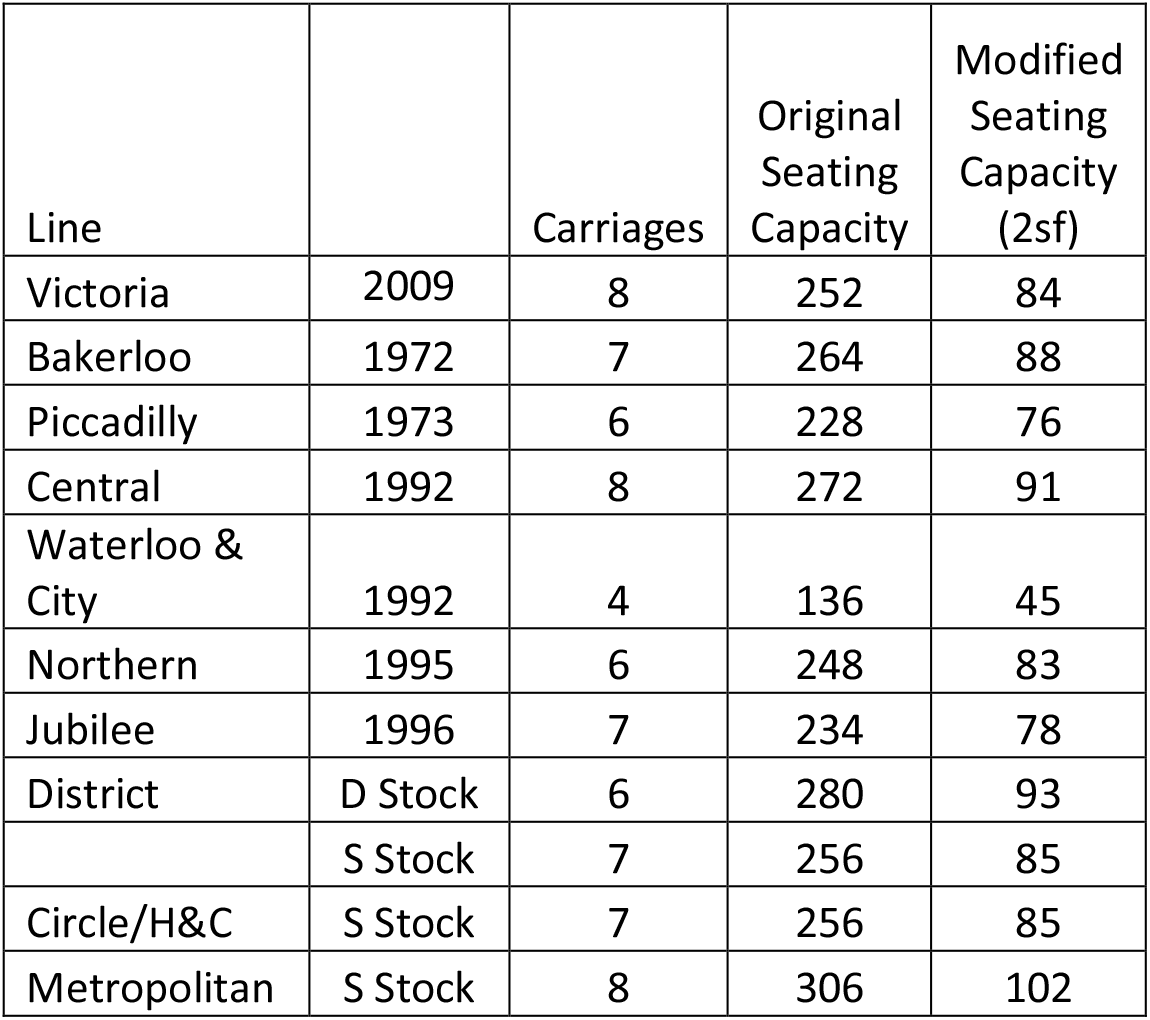
Modified Seating Capacity Resulting from Application of Social Distancing Policy

| Line |  | Carriages | Original Seating Capacity | Modified Seating Capacity (2sf) |
| --- | --- | --- | --- | --- |
| Victoria | 2009 | 8 | 252 | 84 |
| Bakerloo | 1972 | 7 | 264 | 88 |
| Piccadilly | 1973 | 6 | 228 | 76 |
| Central | 1992 | 8 | 272 | 91 |
| Waterloo & City | 1992 | 4 | 136 | 45 |
| Northern | 1995 | 6 | 248 | 83 |
| Jubilee | 1996 | 7 | 234 | 78 |
| District | D Stock | 6 | 280 | 93 |
|  | S Stock | 7 | 256 | 85 |
| Circle/H&C | S Stock | 7 | 256 | 85 |
| Metropolitan | S Stock | 8 | 306 | 102 |

### Estimating the Interaction Between Seating and Standing Capacity

The final determination is the interaction between the seating and standing capacities. This required a move away from the generalisations in the previous two estimations and incorporation of the geometric properties of the transport.

#### Worked Example – Victoria Line Trailer Car

It is clear that standing cannot occur between the seated areas if the seated areas are at capacity (i.e. 2 in each row). Therefore the area between the seats must be excluded from the seating capacity calculations.

Although the person density has been used to revise the standing capacity figures, there are other factors that play an over-riding role. The end seats in the seated areas contain a glass partition and this allows for standing positions standing close to the glass partition such that there is a separation of 2 metres between opposing standing positions and this can be reproduced on the opposing side. However the separation between standing and sitting passengers is less but with a physical barrier.

Utilising this new arrangement, the carriage has 4 standing areas. 2 of these areas have a capacity of 2 people and 2 have a capacity of 4 people. Therefore the total in each carriage is 12 people standing for the Victoria Line Trailer Car. The Motor Car has 2 less standing positions as this is replaced by the driver’s carriage.

Therefore for the example of the Victoria Line carriages, there would be a capacity as shown below.

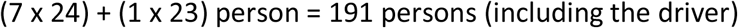

### Consideration of Other Motor and Trailer Cars

The schematics for the remaining carriages were inspected and the same rationale used to populate Table 4 whilst again taking into consideration the geometric properties of the cars as well as the seating arrangements and presence of physical barriers. The analysis proceeded similarly to the worked example above but there were a few important differences.

The following assumptions were used:

1. On the Bakerloo line, the motor car seating arrangement differs from the trailer car in that in addition to the driver compartment there is a section in the middle of the car that has a denser seating arrangement. Given the social distancing guidance, it is assumed that the seating capacity would be equivalent to the Victoria Line trailer carriage as the 2 metre distance precludes more than one person occupying the same vertically aligned seat (vertical being defined here as perpendicular to the length of the carriage). Therefore the seats on each opposing row should be considered as viable for more than one person only in the horizontal direction since only then is a separation of 2 metres possible.
2. The analysis does not consider the use of the uncoupling car and instead focuses solely on the use of the driver and trailer cars.
3. The surface carriages are wider but this does not increase the standing capacity due to the area needed for social distancing.

**Table 4:** Final Total Capacity Calculations for Different Underground Models

| Line |  | Carriages | Modified Capacity |
| --- | --- | --- | --- |
| Victoria | 2009 | 8 | 191 |
| Bakerloo | 1972 | 7 | 167 |
| Piccadilly | 1973 | 6 | 143 |
| Central | 1992 | 8 | 191 |
| Waterloo & City | 1992 | 4 | 95 |
| Northern | 1995 | 6 | 143 |
| Jubilee | 1996 | 7 | 167 |
| District | D Stock | 6 | 193 |
|  | S Stock | 7 | 225 |
| Circle/H&C | S Stock | 7 | 225 |
| Metropolitan | S Stock | 8 | 257 |

## Discussion

The paper makes a number of assumptions based on the schematics of different underground and overground cars on the London Underground. The capacity was calculated on the basis of these assumptions and shows a reduction in the projected capacity of the cars based on the adherence to social distancing principles. However it should be noted that another source gives a figure of 3 persons-per-metre squared as that intended in the original design of the transport cars (What do they know, 2020 (a)). Therefore the original standing capacity in Table 2 is an overestimation of standing capacity by 67%. This in turn means that social distancing person density is a 3800% reduction in standing capacity compared to the intended design although the subsequent analysis incorporating the geometry of the cars allows for a higher standing capacity.

The main findings are that standing can occur in specified portions of the car but should avoid the area between the seats when they are occupied at capacity. The main standing areas would be in the space horizontally (i.e. parallel to the length of the car) adjacent to the seating areas and in front of the doors. These areas are of two types – wide and thin. For the wide areas, four standing people can be accommodated standing in the four corners of the area so as to maximally separate from each other. The passengers can stand close to the windows separating the standing area from the seating area. For the thin areas, two people can be accommodated on opposite sides. There are various seating areas but as a general rule, seating should be considered in the horizontal orientation and each row of seating can accommodate two people.

The reduced capacity of each car requires a compensatory increase in the frequency of the underground trains or else an increase in the cars per train if this is possible, depending on the demand. A mismatch between the train capacity and the passengers may result in crowding on platforms depending on how the flow is managed.

Another consideration is for the arrangements for boarding and alighting from the car which will require an increased time so as to enable passengers to maintain a distance of 2 metres from each other. Passengers would need to communicate with each other about how they will alight from the car whilst maintaining social distance. The standing and seated positions could be marked and an educational campaign initiated to ensure that commuters understand the logistics of this arrangement.

Further consideration will be needed for the use of the glass barriers in separating standing and sitting passengers as if this cannot be used, then it would reduce the standing capacity.

## Data Availability

The data is contained in the tables in the paper.

## Drawbacks

The current paper uses data from an information sheet from 2007 and derived data from 2016 as the starting point for the analysis. There may be more recent data of relevance but this was not identified in the literature and supplemental searches. The modelling does not factor in the effects of forced ventilation, the dissipation of fine water droplets or the integrity of glass partitions which are assumed to offer a barrier between those seated and standing close to the partition. The modelling does not factor in changes to the number of cars that mirror changes in the number of passengers over time. The modelling also does not factor in passengers that require more space.

